# Low muscle strength is associated with cardiovascular autonomic neuropathy in patients with type 2 diabetes

**DOI:** 10.1101/2024.07.16.24310535

**Authors:** Chan-Hee Jung, Yoon Young Cho, Dug-Hyun Choi, Bo-Yeon Kim, Sang-Hee Jung, Chul-Hee Kim, Ji-Oh Mok

## Abstract

**Background:** Several studies have investigated whether sarcopenia is associated with diabetic microvascular complications, but very few have examined associations between sarcopenia and cardiovascular autonomic neuropathy (CAN) in patients with type 2 diabetes mellitus (T2DM). Therefore, we investigated associations of muscle strength (handgrip strength [HGS]) and mass (appendicular skeletal muscle mass index [ASMI]) and CAN in patients with T2DM.

**Methods:** We enrolled 342 T2DM patients (mean age 56.1±11.5 years; 58.2% male) in this retrospective, cross-sectional study. Cardiovascular reflex tests were used to assess CAN according to Ewing’s protocol. Relative HGS was determined after normalizing absolute HGS to body weight (HGS/body weight [kg]). We defined low HGS and low ASMI according to a consensus report of the Asian Group for Sarcopenia. Logistic regression analyses were carried out to assess the associations between relative HGS or ASMI quartiles and the presence of CAN in patients with T2DM.

**Results:** The prevalence rates of CAN, low HGS, and low ASMI in the study subjects were 34.8%, 17.3%, and 18.7%, respectively. Low HGS was significantly more prevalent in patients with CAN than those without CAN (23.5% vs. 13.9%, p=0.025). The CAN scores were significantly and negatively correlated with relative HGS but not with ASMI. Relative HGS was negative correlated with age, glycated hemoglobin, fasting plasma glucose, hsCRP, body mass index, and HOMA-IR and positively correlated with ASMI. The prevalence of CAN gradually increased with decreasing quartile of relative HGS (28.4% in Q4, 31.8% in Q3, 34.2% in Q2, and 45.3% in Q1, p=0.02 for trend). Multivariable-adjusted prevalence ratios (PRs) for CAN, determined by comparing the first, second, and third quartiles with the fourth quartile of relative HGS, were 4.4 with a 95% confidence interval (95% CI) of [1.1 to 17.3]), 2.3 (95% CI [0.8 to 6.9]), and 1.2 (95% CI [0.4 to 3.7]), respectively. The prevalence of CAN and the PRs (95% [CI]) for CAN based on ASMI were not statistically significant.

**Conclusions:** Our findings suggest that low muscle strength rather than low muscle mass was significantly associated with the presence of CAN. Therefore, HGS testing could help identify patients who would benefit from screening for earlier diagnosis of CAN.

## Introduction

Cardiovascular autonomic neuropathy (CAN) refers to impaired autonomic control of the heart and vessels and is a common microvascular complication of diabetes.^1,2^ Numerous studies have reported that CAN is associated with life-threatening clinical entities, such as silent myocardial ischemia, coronary artery disease, peri-operative cardiovascular (CV) instability, and cardiac arrhythmia, resulting in increased cardiovascular and all-cause mortality.^3–7^ Although there is definitive evidence of the prognostic relevance of CAN, it is underdiagnosed and one of the most overlooked complications of diabetes.^1,2^ Timely diagnosis and efforts to overcome under-diagnosis of CAN are important to avoid serious events. One strategy to overcome under-diagnosis of CAN is to select candidates for active screening based on well known risk factors for the development and progression of CAN in patients with T2DM, such as hyperglycemia, hypertension (HTN), dyslipidemia, and obesity; additional risk factors need to be identified to effectively predict CAN.^8–12^

Sarcopenia is defined as the loss of skeletal muscle mass and reduced muscle strength and/or physical performance.^13^ In patients with T2DM, the presence of sarcopenia is receiving growing attention due to its increasing prevalence and the strong negative impact on prognosis.^14^ Previous studies revealed a bidirectional relationship between sarcopenia and diabetic vascular complications.^14–16^ Several studies have reported associations between sarcopenia and other diabetic microvascular complications^17,18^ but, to our knowledge, there have been very few studies investigating the association between sarcopenia and CAN in T2DM.^19,20^

Previous studies have reported subjects with sarcopenia had a greater impairment of cardiac autonomic function and heart rate variability (HRV) than those without sarcopenia.^21,22^ However, most such studies were carried out in obese patients, operated patients, or elderly individuals without DM.^21,22^ Because there have been very few studies evaluating the impact of sarcopenia on CAN in subjects with T2DM, our goal in this study was to investigate the relationships between CAN and two markers of sarcopenia (muscle strength and mass) in Korean patients with T2DM. Association between sarcopenia and CAN could help identify patients who would benefit from active screening for early diagnosis of CAN. Furthermore, interventions to modify sarcopenia may improve the prognosis of patients with CAN.

## Methods

### Study subjects

We conducted a retrospective, cross-sectional study of patients with T2DM who were treated at Soonchunhyang University Bucheon Hospital between April 2020 and July 2023. Among patients who were treated at the Endocrinology and Metabolism Clinic, 342 who underwent handgrip strength measurement and bioelectrical impedance analysis (BIA) and were assessed for CAN were enrolled in this study. The study protocol was approved by the Institutional Review Board (IRB) of Soonchunhyang University School of Medicine, Bucheon Hospital (IRB number: 2024-04-001). The IRB waived the requirement for informed consent owing to the retrospective design of this medical record review study.

### Assessment of muscle strength and muscle mass

Muscle strength was evaluated by HGS testing twice for each hand using a Smedley analog hand dynamometer (No. 04125; MIS, Tokyo, Japan) in a standing position, and the maximum value was used in the analyses. Relative HGS was calculated as absolute HGS (kg) divided by body weight to reduce the impact of body size.^23^ We used bioimpedance analysis (BIA) to assess muscle mass. Appendicular skeletal muscle mass (ASM) was calculated by summing the lean mass in the arms and legs, which primarily represents skeletal muscle mass. The ASM index (ASMI) was calculated as ASM divided by height squared (ASM/Ht^2^; kg/m^2^). According to the consensus report of the Asian Working Group for Sarcopenia, we defined low muscle strength as HGS <26 kg for men and <18 kg for women and defined low muscle mass as ASMI <7 kg/m^2^ in men and <5.7 kg/m^2^ in women.^13^ Subjects were divided into relative HGS quartiles or ASMI quartiles for statistical analysis.

### Diagnosis of CAN

Autonomic function tests (AFT) were performed by a single operator in the morning in a quiet room, and the results were analyzed by one investigator. The subject was instructed to avoid consuming food, coffee, alcohol, and smoking for at least 2 h before the tests. The presence of CAN was assessed using the five standard non-invasive cardiovascular reflex tests (CVRTs) proposed by Ewing and Clarke more than 30 years ago [24] and that are now considered the cornerstone of diagnosis. The five CVRTs are the heart rate (HR) responses to (i) deep breathing (beat-to-beat variation), (ii) standing (30:15 ratio), and (iii) the Valsalva maneuver and the blood pressure (BP) response to (iv) standing and (v) a sustained handgrip. The CVRTs were assessed automatically from electrocardiography recordings using the DICAN evaluation system (Medicore Co., Ltd., Seoul, Korea). Each CVRT was assigned a score of 0, 0.5, or 1 depending on whether the result was normal, borderline, or abnormal, respectively as previously described by Bellavere et al.^25^ The presence of CAN was quantified by summing the scores of the five tests, and was defined as the presence of at least two abnormal tests or an autonomic neuropathy score ≥2.

### Other covariates

Height and weight were measured to the nearest 0.1 cm and 0.1 kg, respectively. Body mass index (BMI) was calculated as body weight (kg) divided by height squared (meters). Blood samples were collected after overnight fasting. Glycated hemoglobin (HbA1c) was measured by ion-exchange high-performance liquid chromatography (Bio-Rad, Hercules, CA, USA) using a methodology aligned with the Diabetes Control and Complications Trial and National Glycohemoglobin Standardization Program standards. Estimated glomerular filtration rate (eGFR) was calculated using the Modification of Diet in Renal Disease study equation, and the urine albumin-to-creatinine ratio (ACR) was assessed in spot urine specimens. Insulin resistance (IR) status was evaluated using the formula for the homeostasis model assessment–insulin resistance (HOMA-IR) index: (fasting insulin [µIU/mL] × fasting plasma glucose [mmol/L])/22.5.

### Statistical analysis

Data are reported as mean ± standard deviation (SD) or median (interquartile range) for continuous variables or as number of subjects (percentage) for categorical variables. The baseline characteristics of the study subjects according to the presence of CAN were compared using two-sample independent *t* tests for continuous variables and Chi-square tests for categorical variables as appropriate. To evaluate linear trends in the prevalence of CAN according to relative HGS and ASMI quartiles, *p* values were calculated using linear-by-linear association tests for categorical variables. Correlations between relative HGS, ASMI, and CAN score were assessed using the Spearman’s rank correlation coefficient. Multivariate logistic regression analyses were performed to estimate the independent effects of relative HGS or ASMI quartile on the prevalence ratios (PRs) of CAN after adjustment for possible confounding factors (age, sex, BMI, current alcohol drinking, current smoking, duration of diabetes, HTN, HbA1c, triglyceride (TG), low density lipoprotein-cholesterol (LDL-C), eGFR, ACR, high-sensitivity c-reactive protein (hsCRP), HOMA-IR, beta-blockers, angiotensin-receptor blockers (ARBs), sodium-glucose cotransporter 2 inhibitors (SGLT2is), and insulin treatment). Skewed variables (hsCRP, TG, and ACR) were transformed logarithmically to improve normality before analyses. P values less than 0.05 were considered significant. All statistical analyses were performed using SPSS 19.0 statistical software (IBM Corp., Armonk, NY, USA).

## Results

### General characteristics of the subjects

The general characteristics of the study subjects are presented in Table 1. A total of 342 Korean T2DM patients (199 male, 143 female, mean age 56.1±11.5 years) was enrolled in this retrospective, cross-sectional study. The CAN prevalence was 34.8% (n=119), and the mean BMI and duration of DM were 25.7 kg/m^2^ and 7.0 years, respectively. The mean HbA1c of the study population was 8.5%, and the proportions of subjects treated with oral hypoglycemic agents (OHA) or insulin were 65.2% and 15.8%, respectively. The mean HGS and ASMI were 29.7 kg and 7.3 kg/m^2^, respectively, and the proportion (number) of patients with low HGS and low ASMI were 17.3% (n=59) and 18.7% (n=64).

**Table 1.** General characteristics of study patients according to the presence of CAN.

|  | Total | CAN (–) | CAN (+) | P-value |
| --- | --- | --- | --- | --- |
| Frequency, n (%) | 342 | 223 (65.2%) | 119 (34.8%) |  |
| Age (years) | 56.1±11.5 | 55.4±11.6 | 57.4±11.3 | 0.123 |
| Duration of diabetes | 7.0±7.5 | 6.6±7.5 | 7.9±7.5 | 0.149 |
| Sex (men), n (%) | 199 (58.2%) | 137 (61.4%) | 62 (52.1%) | 0.096 |
| HbA1C (mmol/mol) | 69.4±2.0 | 67.2±1.0 | 72.7±3.0 | 0.070 |
| FPG (mmol/L) | 8.42±2.89 | 8.26±2.73 | 8.72±3.15 | 0.185 |
| Total cholesterol (mg/dL) | 159±47 | 156±45 | 164±51 | 0.140 |
| TG (mg/dL) | 125 (90–178) | 122 (90–173) | 127 (91–195) | 0.191 |
| HDL-C (mg/dL) | 47±11 | 47±11 | 47±11 | 0.778 |
| LDL-C (mg/dL) | 87±35 | 85±34 | 90±37 | 0.191 |
| HTN, n (%) | 119 (34.8%) | 74 (33.6%) | 45 (40.2%) | 0.240 |
| hsCRP (mg/L) | 0.08 (0.04–0.22) | 0.08 (0.04–0.23) | 0.08 (0.04–0.21) | 0.593 |
| ACR | 1.2 (0.6–3.7) | 1.1 (0.6–3.2) | 1.4 (0.6–6.2) | 0.905 |
| eGFR | 90.1±17.0 | 90.6±17.4 | 87.2±20 | 0.096 |
| HOMA-IR | 2.9 (1.7–4.8) | 2.8 (1.6–4.4) | 3.2 (1.8–5.1) | 0.095 |
| BMI | 25.7±4.0 | 25.7±3.9 | 25.8±4.1 | 0.703 |
| HGS (kg) | 29.7±9.6 | 30.8±9.4 | 27.6±9.6 | 0.004 |
| Relative HGS | 0.43±0.12 | 0.44±0.11 | 0.40±0.12 | 0.006 |
| ASMI (kg/m<sup>2</sup>) | 7.3±1.0 | 7.4±1.1 | 7.2±1.0 | 0.164 |
| Low HGS, n (%) | 59 (17.3%) | 31 (13.9%) | 28 (23.5%) | 0.025 |
| Low ASMI, n (%) | 64 (18.7%) | 44 (19.7%) | 20 (16.8%) | 0.509 |
| Smoking, n (%) | 89 (26%) | 63 (28.3%) | 26 (21.8%) | 0.328 |
| Alcohol, n (%) | 110 (32.2%) | 69 (30.9%) | 41 (34.5%) | 0.455 |
| Diabetic retinopathy, n (%) | 64 (25.4%) | 37 (22.8%) | 27 (30%) | 0.211 |
| DPN, n (%) | 112 (41.8%) | 66 (37.9%) | 46 (48.9%) | 0.081 |
| OHA, n (%) | 223 (65.2%) | 149 (68%) | 74 (66.1%) | 0.718 |
| Metformin, n (%) | 213 (62.3%) | 145 (65%) | 68 (57.1%) | 0.077 |
| DPP4I, n (%) | 132 (38.6%) | 82 (36.8%) | 50 (42%) | 0.026 |
| SGLT2I n, (%) | 48 (14%) | 39 (17.5%) | 9 (7.6%) | 0.004 |
| GLP1RA n, (%) | 17 (5%) | 12 (5.4%) | 5 (4.2%) | 0.056 |
| Insulin, n (%) | 54 (15.8%) | 34 (15.2%) | 20 (16.8%) | 0.051 |
| ARB | 97 (28.4%) | 61 (27.4%) | 36 (30.3%) | 0.042 |
| Beta blockers | 16 (4.7%) | 14 (6.3%) | 2 (1.7%) | 0.011 |

Data are reported as mean ± standard deviation (SD) or median (interquartile range) or as number of subjects (percentage).
ACR, Albumin/creatinine ratio; ARB, angiotensin receptor blocker; ASMI, appendicular skeletal muscle index; BMI, body mass index; CAN, cardiovascular autonomic neuropathy; DPN, diabetic peripheral neuropathy; DPP4I, dipeptidyl peptidase-4 inhibitor; FPG, fasting plasma glucose; GLP1RA, glucagon like peptide-1 receptor agonist; HbA1C, glycated hemoglobin; HDL-C, high density lipoprotein-cholesterol; HGS, handgrip strength; HOMA-IR, homeostasis model assessment–insulin resistance; hsCRP, high-sensitivity c-reactive protein; LDL-C, low density lipoprotein-cholesterol; OHA, oral hypoglycemic agent; SGLT2I, sodium-glucose cotransporter 2 inhibitor; TG, triglyceride

### Comparisons of clinical characteristics according to the presence or absence of CAN

The general characteristics of subjects according to the presence or absence of CAN are presented in Table 1. The mean HGS and relative HGS were lower in subjects with CAN than in subjects without CAN (27.6 kg vs. 30.8 kg, p=0.004, 0.40 vs. 0.44, p=0.006, respectively). A greater proportion of subjects with CAN had low HGS than those without CAN (23.5% vs. 13.9%, p=0.025). There were no significant differences in mean age, duration of DM, BMI, HTN, glucose parameters, lipid profiles, eGFR, ACR, hsCRP, and HOMA-IR according to the presence or absence of CAN. In addition, there were no significant differences in treatment modalities such as insulin or OHA between subjects with CAN and those without CAN. The proportion of subjects using SGLT2is or beta-blockers was significantly larger in subjects without CAN than those with CAN (17.5% vs. 7.6%, p=0.004, 6.3% vs. 1.7, p=0.011, respectively), and use of ARB was more prevalent in those with CAN than those without CAN (30.3 % vs. 27.4%, p=0.042).

### Prevalence of CAN according to relative HGS and ASMI quartiles

The prevalence of CAN according to relative HGS and ASMI quartiles is shown in Figure 1. The prevalence declined significantly and progressively from the lowest to highest quartiles of relative HGS (45.3%, 34.2%, 31.8%, and 28.4%, respectively, p=0.02 for trend; Fig. 1[a]). On the other hand, the prevalence of CAN did not differ from the lowest to highest quartiles of ASMI (36.8%, 35.2%, 34.5%, and 33%, respectively, p=0.197 for trend; Fig. 1[b]).

**Fig. 1.**
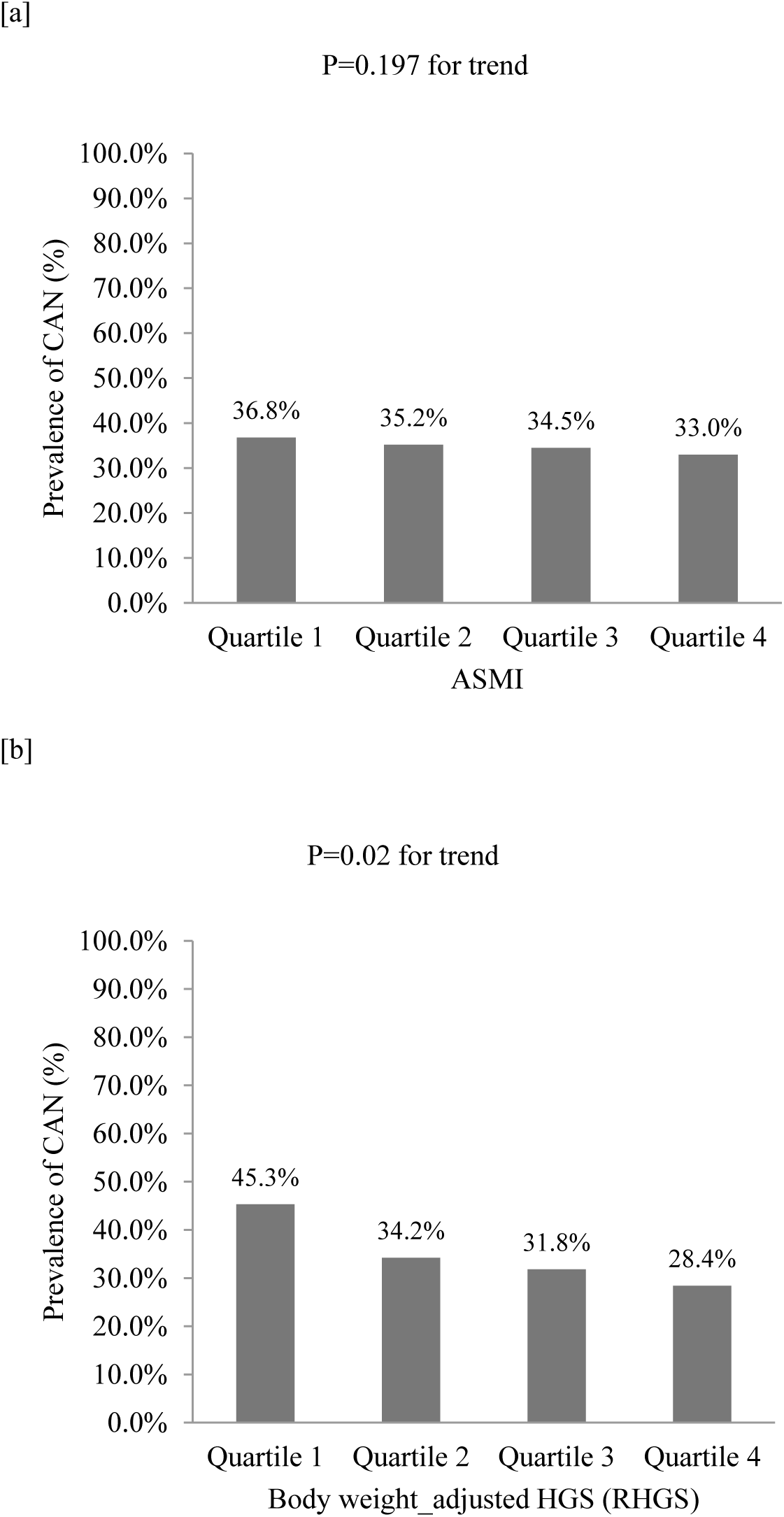
The prevalence of CAN in study patients according to quartile of relative HGS or ASMI.

### Bivariate correlations of CAN score and relative HGS and ASMI with clinical variables

Correlations of CAN score, relative HGS, and ASMI with clinical variables are shown in Table 2. The CAN score showed a significant negative correlation with relative HGS (r=−0.137, p=0.012). However, there was no significant correlation between CAN score and ASMI. Relative HGS showed significant negative correlations with age, FBS, HbA1c, hsCRP, BMI, and HOMA-IR (r=-0.116, p=0.032; r=-0.132, p=0.015; r=-0.119, p=0.03; r=-0.201, p=0.003; r=-0.387, p<0.001; and r=-0.198, p=0.001, respectively) and a positive correlation with ASMI (r=0.334, p<0.001). The ASMI was negatively correlated with age and HDL-C (r=-0.403, p<0.001; r=-0.172, p=0.002, respectively) and positively correlated with TG, eGFR, BMI, and HOMA-IR (r=0.192, p<0.001; r=0.158, p=0.003; r=0.439, p<0.001; and r=0.12, p=0.049, respectively).

**Table 2.** Correlation analyses for relative HGS, ASMI, and CAN score with clinical variables in total subjects.

|  | Relative HGS | ASMI | CAN Score |
| --- | --- | --- | --- |
|  | r (p-value) | R (p-value) | R (p-value) |
| Relative HGS | - | 0.334 (<0.001) | -0.137 (0.012) |
| ASMI | 0.334 (<0.001) | - | -0.051 (0.350) |
| CAN score | -0.137 (0.012) | -0.051 (0.350) | - |
| Age (years) | -0.116 (0.032) | -0.403 (<0.001) | 0.09 (0.097) |
| Duration of diabetes (years) | -0.01 (0.861) | -0.042 (0.445) | 0.061 (0.263) |
| FPG (mg/dL) | -0.132 (0.015) | -0.021 (0.705) | 0.023 (0.668) |
| HbA1c (%) | -0.119 (0.03) | -0.023 (0.675) | 0.102 (0.064) |
| TC (mg/dL) | -0.06 (0.272) | -0.014 (0.790) | 0.006 (0.910) |
| TG (mg/dL) | -0.103 (0.057) | 0.192 (<0.001) | -0.033 (0.542) |
| HDL-C (mg/dL) | -0.026 (0.640) | -0.172 (0.002) | 0.07 (0.215) |
| LDL-C (mg/dL) | -0.041 (0.461) | 0.021 (0.707) | 0.008 (0.881) |
| hsCRP (mg/L) | -0.201 (0.003) | 0.047 (0.486) | -0.02 (0.770) |
| ACR | -0.108 (0.071) | 0.017 (0.780) | 0.03 (0.620) |
| eGFR | 0.036 (0.513) | 0.158 (0.003) | -0.089 (0.101) |
| BMI | -0.387 (<0.001) | 0.439 (<0.001) | -0.003 (0.959) |
| HOMA-IR | -0.198 (0.001) | 0.12 (0.049) | 0.027 (0.659) |

ACR, Albumin/creatinine ratio; ASMI, appendicular skeletal muscle index; BMI, body mass index; CAN, cardiovascular autonomic neuropathy; FPG, fasting plasma glucose; HbA1C, glycated hemoglobin; HDL-C, high density lipoprotein-cholesterol; HGS, handgrip strength; HOMA-IR, homeostasis model assessment–insulin resistance; hsCRP, high-sensitivity c-reactive protein; LDL-C, low density lipoprotein-cholesterol; TG, triglyceride

### Associations between CAN and relative HGS or ASMI quartile

Multivariate logistic regression analysis adjusted for confounding variables was used to evaluate the associations between HGS, ASMI, and CAN (Table 3). When the highest relative HGS quartile group was used as a reference in the univariate model, patients in the lowest quartile (Q1) were 2.1-fold more likely to have CAN (OR 2.1; 95% CI [1.1 to 3.9]). This significant association for group Q1 remained even after adjusting for age, duration of DM, HTN, smoking, alcohol, BMI, HbA1c, eGFR, ACR, TC, hsCRP, HOMA-IR, insulin treatment, SGLT2is, beta-blockers, and ARBs (OR 4.4, 95% CI [1.1 to 17.3]). However, there were no significant association between ASMI and CAN in either the unadjusted model or after multivariate adjustment.

**Table 3.** Multiple logistic regression analysis of the association of sarcopenia with presence of CAN.

|  | Model 1 |  | Model 2 |  |
| --- | --- | --- | --- | --- |
|  | PRs (95% CI) | P-value | PRs (95% CI) | P-value |
| Relative HGS |  |  |  |  |
| Quartile 1 | 2.1 [1.1, 3.9] | 0.021 | 4.4 [1.1, 17.3] | 0.033 |
| Quartile 2 | 1.3 [0.7, 2.5] | 0.422 | 2.3 [0.8, 6.9] | 0.126 |
| Quartile 3 | 1.2 [0.6, 2.2] | 0.622 | 1.2 [0.4, 3.7] | 0.731 |
| Quartile 4 | 1 [Reference] |  | 1 [Reference] |  |
| ASMI |  |  |  |  |
| Quartile 1 | 1.2 [0.6, 2.2] | 0.601 | 1.4 [0.3, 6.7] | 0.648 |
| Quartile 2 | 1.1 [0.6, 2.1] | 0.75 | 1.2 [0.3, 5.3] | 0.775 |
| Quartile 3 | 1.1 [0.6, 2.0] | 0.831 | 1.1 [0.3, 4.0] | 0.835 |
| Quartile 4 | 1 [Reference] |  | 1 [Reference] |  |

## Discussion

We demonstrated that T2DM patients with low HGS are at a higher risk for the presence of CAN; however, low ASMI was not associated with the presence of CAN. These findings suggest that assessment of muscle strength using HGS could be invaluable in clinical practice to identify high-risk T2DM subjects who require earlier and active screening for CAN.

There is considerable variation in the reported prevalence of CAN, depending upon the diagnostic criteria, test tool, interpretation of results, and study population.^2^ The prevalence of clinical and subclinical CAN has been estimated to be 20–65% of diabetic patients, which is greater than the prevalence of other diabetic complications. ^2,26^ There is definitive evidence of the prognostic relevance of CAN for all-cause and CV mortality, and the risks are increased even at an asymptomatic stage.^27^ Despite the importance of early detection of CAN, there are several possible barriers to testing: there is no harmonized definition of CAN, there is a wide variety of tools for assessing CAN, testing can be time-consuming and cumbersome, the results may be difficult to interpret, specific treatments for CAN are lacking, and testing may be considered unnecessary in asymptomatic cases.^28^ Despite these barriers to testing, screening for CAN using Ewing’s tests and HRV may allow diagnosis of at an earlier stage where risk factor modification is possible and reversibility is more likely. The established risk factors for development and progression of CAN are age, duration of diabetes, glycemic control, JTN, dyslipidemia, obesity, and smoking.^9,11,29^ In addition, some studies have suggested glycemic variability, vitamin D, oxidative stress, and chronic inflammation as additional risk factors for CAN.^30–32^

There are a few reports in the literature correlating sarcopenia with cardiac autonomic modulation. Early studies suggested that low muscle mass and muscle strength lead to reduced parasympathetic modulation and may be associated with abnormal autonomic modulation. A cross-sectional study of 76 elderly Brazilians showed that greater impairment of HRV was evident in subjects with sarcopenia than in those without sarcopenia.^22^ In the study, muscular strength was assessed using HGS, muscle mass was assessed using the skeletal muscle index, and cardiac autonomic modulation was evaluated using a heart rate (HR) monitor to record five minutes of successive RR intervals. The analysis of the RR-interval variability was carried out in time and frequency domains. The non-sarcopenia group was that with adequate muscle strength and physical performance but insufficient muscle mass or adequate muscle mass. Sarcopenia was defined as insufficient muscle mass plus insufficient muscle strength and/or poor physical performance. The parameters of autonomic heart rate modulation differed significantly between elderly sarcopenic and non-sarcopenic participants. Elderly sarcopenic participants exhibited lower parasympathetic-associated modulation. Baek et al. investigated the relationship between overweight with low muscle mass and cardiac autonomic function as assessed by HRV in a five-min ECG recording in 1,150 healthy Korean workers.^21^ The overweight group showed reduced parasympathetic modulation compared with the non-overweight group. In addition, the overweight-with-low-muscle-mass group had a lower HRV than the overweight-with-high-muscle-mass group.

Hyperinsulinemia and IR can induce the development of a sympathovagal imbalance with SNS predominance^33^; hyperinsulinemia-induced SNS activation may then result in further IR and hyperinsulinemia. Weight loss using lifestyle interventions and bariatric surgery may help improve cardiac autonomic function by reducing IR and hyperinsulinemia, leading to a reduction in SNS activation. Several studies have reported that weight reduction through bariatric surgery (BS) improves cardiac autonomic modulation.^34,35^ Furthermore, the presence of low muscle mass and/or strength in the preoperative period has been shown to attenuate the improvement in cardiac autonomic function after BS compared with patients without sarcopenia.^36^ A recent study by Carvalho et al. showed that women who underwent BS experienced less of an improvement of HRV during the follow-up period among women with low muscle mass and /or handgrip strength.^36^

To the best of our knowledge, there have been few studies assessing associations of sarcopenia and CAN in T2DM, and the findings were not consistent with those of our study.^19,20^ The primary aim of the study by Mikura et al. was to investigate the association between sarcopenia and severity of diabetic peripheral neuropathy (DPN) in 261 Japanese patients (median age; 67 years, median duration of DM; 10 years).^19^ They also evaluated the association of autonomic neuropathy and sarcopenia. The coefficient of variation of the R-R interval was assessed using ECG as a marker of autonomic function, and autonomic neuropathy was noted if CV R-R was lower than the reference ranges. In addition, sarcopenia was identified based on the AWGS 2019 diagnostic criteria, consistent with our current study. The Mikura et al. study showed that neither autonomic neuropathy nor DPN was significantly correlated with sarcopenia. Another study by Ozgur Y et al. examined the relationships between several diabetic neuropathy subtypes and low muscle mass/low muscle strength in a cross-sectional study of 602 diabetic patients (mean age; 60.2 years).^20^ In the study, diabetic neuropathy was classified into subtypes of diabetic sensorimotor polyneuropathy or diabetic autonomic neuropathies (DAN), and DAN was further subclassified into four subgroups of gastrointestinal autonomic neuropathy, genitourinary autonomic neuropathy, sudomotor autonomic neuropathy, and CAN. The Ozgur study used HR while sitting, HR response to standing, and BP response to standing to diagnose CAN in the presence of abnormality in two of three tests. When the DAN sub-groups were considered, the prevalence of CAN did not differ significantly between the sarcopenic and non-sarcopenic groups. In addition, there was no significant association between CAN and low muscle mass or low muscle strength in logistic regression analyses. Specifically, the prevalence of diabetic neuropathy in the low muscle strength group was greater than in the low muscle mass group. They found that although the skeletal muscle mass index was higher in low muscle strength when compared to low muscle mass group, their muscle qualities were much lower in low muscle strength. The latter finding suggests that loss of muscle strength is more important than loss of muscle mass in terms of the prognosis of diabetic neuropathy. In our study, subjects with low HGS had a greater CAN prevalence than subjects with low ASMI (47.5% vs. 31.2%, data not shown). The prevalence of CAN increased progressively with decreasing quartile of relative HGS, and the lowest quartile of relative HGS was significantly associated with the PR of CAN. However, the prevalence of CAN and PRs for CAN based on ASMI were not significant. Our findings study also suggest that muscle strength may be a more useful diagnostic marker of the presence of CAN than muscle mass.

Most previous studies used HRV derived from ECG recordings as a diagnostic tool for assessment of CAN, whereas there have been no studies investigating the correlation of CAN score based on CVRTs as assessed by Ewing’s test and sarcopenia parameters. Although agreement between indices of HRV derived from ECGs and CVRTs has been reported, CVRTs are the gold standard for the diagnosis of CAN.^37^ We analyzed the correlations of relative HGS, ASMI, and CAN score with clinical parameters and found that relative HGS was significantly and negatively correlated with hsCRP, BMI, HOMA-IR, and glucose parameters. The significant correlations of relative HGS with inflammatory markers, obesity, IR index, and glycemia, all of which are risk factors of CAN, indicate that they contribute to the association between muscle strength and presence of CAN. On the other hand, ASMI was marginally and positively correlated with HOMA-IR, but there was no correlation with hsCRP and glucose parameters. Overall, the CAN score was significantly correlated only with relative HGS not with ASMI. In a Danish multicenter cross-sectional study of T2DM patients, CAN was independently associated with high BP, BMI, and smoking.^26^ However, in our study, the CAN score was not significantly correlated with duration of diabetes, HbA1c, lipid profile, hsCRP, eGFR, ACR, BMI, or HOMA-IR. Previous studies also reported that age, duration of diabetes, eGFR, BP, lipid profile, and HbA1c did not differ between subjects with and without CAN.^30^

Among sarcopenia parameters, muscle strength may represent muscle quality or function. Several studies have reported that low muscle strength, but not muscle mass, is associated with poor physical function and higher mortality.^23,38^, and that muscle strength more strongly influences various diabetic complications than does muscle mass in patients with T2DM.^17^ These findings suggest that measuring muscle mass alone may not be sufficient to predict and protect against DM complications, and preservation of muscle strength may be more important. A previous joint international longitudinal study suggested that HGS is a useful predictor of the risk of CVD and mortality.^23^ In addition, a subanalysis of the ORIGIN trial showed HGS to be associated with CVD mortality in patients with T2DM.^38^

The mechanisms underlying the association between sarcopenia and cardiac autonomic dysfunction are unclear, but some explanations can be suggested. First, CAN and sarcopenia have multifactorial etiologies and common risk factors. Hyperinsulinemia or IR results in early CAN through the development of a sympathovagal imbalance with SNS activation.^33,39^, while IR has also been suggested as a main underlying mechanism of sarcopenia. In addition, increased low-grade chronic inflammation is a common feature in patients with sarcopenia and has been associated with decline in muscle mass and strength and reduced HRV.^40^ Second, increased reactive oxygen species and mitochondrial dysfunction can lead to muscle wasting and autonomic dysfunction.^41,42^ Third, the myokine irisin produced by skeletal muscle may serve as an important cross-organ messenger linking skeletal muscle with the brain, adipose tissue, and CV system to integrate energy expenditure with CV activity.^43^ Irisin controls HR, modulates vagal tone, and promotes CV protection.^44^ Previous studies showed a positive correlation between irisin and HGS in a multivariate analysis.^45,46^ Irisin has been reported to be strongly associated with muscle strength and was recently suggested to be a candidate factor to explain the relationship between muscle strength and parasympathetic tone to the heart, promoting a cardioprotective role.^43,44^ However, since serum irisin was not measured in our study, a direct causal relationship among irisin, muscle strength, and autonomic modulation in patients with T2DM needs to be further examined.

While there is no conclusive evidence of a successful disease-modifying therapy, multifactorial interventions including glycemic control and weight loss might be effective. In addition, beta blockers and renin-angiotensin-aldosterone system inhibitors are known to have a positive effect on autonomic dysfunction, and emerging data on SGLT2i suggest a modulatory effect on cardiovascular autonomic function.^47,48^ In our current study, adjustment of the models for these medications did not substantially change the results.

One strength of the present study is that, to the best of our knowledge, it is the first to investigate directly the relationship between HGS and CAN using CVRTs in patients with T2DM. This study also has some limitations. First, we excluded patients with no CVRT, BIA, or HGS results, which led to a small sample size and a potential for selection bias. Second, because of the retrospective, cross-sectional nature of this study, we could not determine causality of the relationship between CAN and sarcopenia. Future prospective or interventional studies are needed to investigate the longitudinal associations among HGS, ASMI, and CAN in a large sample size.

In conclusion, we demonstrated that HGS was associated with the presence of CAN in Korean patients with T2DM. We also found that ASMI was not associated with the presence of CAN. Low muscle strength is an important cause of morbidity in T2DM and CAN is also a serious complication. In patients with both low muscle strength and CAN, poor quality of life may be expected. Therefore, low muscle strength may be recognized with a simple HGS test performed in clinical practice, allowing early diagnosis of CAN.

## Author contributions

CHJ and JOM contributed to the conception and design of the study; CHJ, YYC, DHC, BYK, and CHK contributed to data collection and management; CHJ, YYC, DHC, BYK, SHJ, CHK, and JOM contributed to acquisition, analysis, or interpretation of the data; CHJ and SHJ contributed to writing the manuscript; JOM contributed to manuscript revision. All authors read and approved the final manuscript.

## Data Availability

The data shown in this article are available from the corresponding author upon a reasonable request.

## Acknowledgements

This study was supported by a grant from Soonchunhyang University.

## Disclosures

None.

## Conflicts of interest

The authors have no conflicts of interest to declare.

## Ethical statement

The study was approved by the Institutional Review Board of Soonchunhyang University Bucheon Hospital.

## Informed consent

Not applicable

## Notes

### Competing Interest Statement

The authors have declared no competing interest.

